# Private Equity Acquisition in Ophthalmology and Optometry: A Time Series Analysis of the Pre-COVID, COVID Pre-Vaccine, and COVID Post-Vaccine Eras

**DOI:** 10.1101/2022.02.03.22270390

**Authors:** Sachi A. Patil, Daniel G. Vail, Jacob T. Cox, Evan M. Chen, Prithvi Mruthyunjaya, James C. Tsai, Ravi Parikh

## Abstract

**Objective:** To identify temporal and geographic trends in private equity (PE) backed acquisitions of ophthalmology and optometry practices in the United States from 2012 to 2021.

**Design:** Cross-sectional time series analysis using acquisition data from 10/21/2019 to 9/1/2021 compared to previously published data from 1/1/2012-10/20/2019. Acquisition data was compiled from 6 financial databases, 5 industry news outlets, and publicly available press releases. Linear regression models were used to compare rates of acquisition.

**Subjects:** 245 PE acquisitions of ophthalmology and optometry practices in the United States between 10/21/2019 and 9/1/2021.

**Measures:** Number of total acquisitions, practice type, locations, provider details, and geographic footprint.

**Results:** 245 practices associated with 614 clinical locations and 948 ophthalmologists or optometrists were acquired by 30 PE-backed platform companies. 18 of 30 platform companies were new compared to our prior study. Of these acquisitions from 10/21/2019 - 9/1/2021, 127, 29, and 89 were comprehensive, retina, and optometry practices, respectively. From 2012 to 2021, monthly acquisitions increased by 0.947 acquisitions per year (p<0.001*). Texas, Florida, Michigan, and New Jersey were the states with the greatest number of PE acquisitions with 55, 48, 29, 28 clinic acquisitions, respectively. Average monthly PE acquisitions were 5.71 per month from 1/1/2019 - 2/29/2020 (pre-COVID), 5.30 per month 3/1/2020-12/31/2020 (COVID pre-vaccine, p=0.8072), and were 8.78/month 1/1/2021-9/1/2021 (COVID post-vaccine, p=0.1971).

**Conclusion:** PE acquisitions increased from 2012-2021 as companies continue to utilize both regionally focused and multi-state models of add-on acquisitions.

## Introduction

Private equity (PE) investment is a major driving force in healthcare today, offering consolidation and the ability to scale up independent practices to compete in a dynamic market. For example, one recent study found that in Maryland and New Jersey, more than 25% of urologists were employed by a private equity backed platform company[1]. However, recent trend of private equity (PE) backed consolidation in ophthalmology and the healthcare system as a whole continues to raise concerns regarding patient care, physician autonomy, and overall health system costs[2, 3]. In the typical PE model, firms invest into an ophthalmic management group, or platform company, which provides streamlined operational efficiency, revenue enhancement, and increased market share [4]. Subsequently, there is an anticipated return on investment within three to six years through recapitalization. PE continues to affect practices in many subspecialities including radiology, dermatology, gastroenterology, and others as they continue to expand their presence[2, 5, 6].

We have previously reported on the rapid increase in private equity investments within the field of ophthalmology and optometry from 2012 through much of 2019 as well as the recapitalization of some of those platform companies [7]. Ophthalmology remains lucrative for the private investment model for several reasons. Firstly, there is a very defined referral path through which the business of the ophthalmologist is clearly defined[8]. Further, high procedure volume with numerous patients, unique pay structures, and an aging population with heavy demand for ophthalmic care creates an ideal market for business growth. Even further, ophthalmologic clinics may potentially have lucrative cash-pay procedures (e.g., cosmetic injections or premium intraocular lenses)[9]. Private equity (PE) investment provides an attractive alternative to hospital-physician alignment, and many specialties have seen a recent rise in PE-driven establishment of consolidated platform companies.

COVID-19 has had a dramatic impact on healthcare operations in the outpatient setting with some studies noting ophthalmology had the highest drop in outpatient volume[10].To prepare for the influx of infected persons, health care facilities largely rescheduled elective or less urgent health care procedures[11]. Given reduced revenue in the setting of the pandemic, there have been furloughs, salary reductions, and loss of benefits for health care workers[12]. On one hand, this loss of revenue may make PE acquisitions slow as the return on investment (ROI) could potentially be less than previously expected. However, the financial pressure on physicians and clinics may also lead to easier acquisition of newly distressed assets. Further, the economic policy of low interest rates since the pandemic began would allow highly capitalized PE firms to more easily obtain financing and expand their footprint[13]. Researchers have speculated that private equity investment model, which mitigates physicians’ personal risk, would be an enticing prospect for healthcare institutions wishing to protect themselves from economic downfall [14]. Anecdotally, physicians in practice have noted a desire to use PE as a lucrative exit from the burdensome tasks of administering their own practices and focusing solely on patient care but employees with limited to no operational control [8].

Therefore, the objective of this study was to assess rates and geographic trends of PE acquisitions of ophthalmology and optometry practices in the United States over the last decade.

## Methods

### Identification of Private Equity Backed Acquisitions

We described the temporal and geographic trends in PE development of platform companies and add-on acquisitions. Of note, the data from January 1^st^, 2012 to October 20^th^, 2019 was previously published by our group[7]. We chose to include this data to provide context and details about private equity over the last decade in the United States. We also characterized PE-acquired practices including description of clinical locations, providers, and presence of ancillary services such as optical shops. Within the October 21^st^, 2019 to September 1^st^, 2021 time frame, there were several subdivisions of interest which were used to guide our analysis. On March 11^th^, 2020, the World Health Organization declared the novel coronavirus a global pandemic[15]. Thus, we compared the periods just prior to the COVID-19 pandemic to the height of the COVID-19 pandemic before vaccinations, or until December 31^st^, 2020. We also compared the period after COVID-19 vaccines were made available to American adults (officially December 14^th^, 2020) and we estimated that this would not have had a sizeable effect until January 2021[16].

Methods for identification and inclusion of private equity backed acquisitions have been described elsewhere[7]. In summary, after the initial investment of creation of a platform company or acquisition of a platform practice group, the platform company will acquire additional practices. We sought to identify and characterize these acquisitions as well as the creation of platform companies, when possible. Financial databases were searched for acquisitions between October 21^st^, 2019 and September 1^st^, 2021. Acquisitions were excluded from this study if the acquisition was of a biotechnology or pharmaceutical company and if the acquired practice had provided services for multiple medical specialties. Transactions involving bankruptcy of the acquired practice or with no identifiable buyer were also excluded. From each acquisition, we collected the following characteristics: name, number of clinical locations, addresses of clinic locations, when possible, number of providers, number of optical shops, and number of practice-owned surgical centers. This study received approval from the Institutional Review Board of Yale University and is compliant with the tenets of the Declaration of Helsinki.

Clinical sites were defined as all practice locations with at least 1 ophthalmologist or optometrist providing clinical care that were not surgical centers, vision rehabilitation centers, or optical shops. A surgical center was defined as any practice location capable of performing laser refractive, cataract, or retinal surgery procedures. Surgical centers also included dedicated ambulatory surgery centers owned by the practice. To capture the data most accurately at time of acquisition, we used information from press releases and web archives dated closest to the transaction closure date. Of note, for certain acquisitions, if we were unable to specify the exact locations of acquired practices, they were not included in the geographic map or used to calculate totals by state. However, we did include these details in the number of total practices.

We identified the platform company, investing PE firm, and date of initial PE investment for each platform. For platform company formations associated with multiple acquired practices, each practice was counted as a separate acquisition. In cases where the transaction closure date was unavailable, the date of the earliest report of the acquisition was used as a surrogate. If the explicit formation of a platform company was not evident, we defined the first investment of a PE firm in an eye care practice or previous eye care management group as the formation of a platform. We also stratified each acquisition into 3 primary practice specialties: comprehensive, optometry, or retina, depending on many services provided within the practice. Descriptive data of platform companies were reported with means, medians, ranges, and standard deviations.

### Platform Financing

For all platform companies in this study, we identified PE investment information from the following databases: Capital IQ, CB Insights, Thomson ONE, and Pitchbook[17–20]. We recorded all PE investors when available, though some were unable to be located or were kept private due to non-disclosure agreements. Companies providing debt refinancing were not reported as investors.

### Geographic Acquisition Trends

We identified the addresses of all clinical locations associated with acquisitions through press releases and web archives of practice websites immediately before the date of acquisition. Addresses were utilized to create a geographic map in GraphPad Prism software version 8 (GraphPad Software, San Diego, CA).

### Rate of Clinic Acquisition Analyses

Mean number of acquisitions per month were compared using two-tailed unpaired t-tests. We specified a linear regression model of time series data with robust standard errors including data from the previously published study, from January 1^st^, 2012 to September 1^st^, 2021. We created a graph of acquisitions for the entire data set after calculating a three-month average number of clinics acquired. Three months was chosen to control for variance in month-to-month data. P values of <0.05 were considered statistically significant.

## Results

### Description of Acquisitions

An estimated total of 245 practices associated with 614 clinical locations and 948 ophthalmologists or optometrists were acquired by 30 PE-backed platform companies (Table 1). Among acquisitions of ophthalmology practices, 23 platform companies acquired 156 practices associated with 510 clinical locations, 548 ophthalmologists, and 243 optometrists. The median (range, SD) number of ophthalmologists and optometrists associated with each ophthalmology acquisition was 3 (0-50, 7.0) and 1 (0-25, 4.1). Each practice was associated with a median of 2 (1-95, 10.6) clinical locations, 0 (0-2, 0.6) surgical centers, 0(0-7, 1.2) optical shops.

Seven platform companies acquired 89 optometry practices associated with 104 clinical locations, 10 ophthalmologists, 147 optometrists. Each practice was associated with a median of 1 (1-19, 3.2) clinical locations, 0(0-1, 0.15) surgical centers and 1 (0-5, 1.2) optical shops.

### Acquisition Trends

The three-month moving average of practice acquisition increased from 2012-2021 (Figure 1). From 2012 to 2021, on average, monthly acquisitions increased by 0.947/year (p<0.001*). During the period from 3/1/2020-12/31/2020 (COVID pre-vaccine), acquisitions decreased by 1.291/month (p=0.295). From 1/1/2021-9/1/2021 (COVID post, the period following introduction of the COVID19 vaccine, acquisitions increased by 2.531/month (p=0.091). The absolute number of monthly acquisitions increased from 2012-2021, with a decline in the absolute number of acquisitions in 2020 and subsequent increase in 2021 (Figure 2).

**Figure 1:**
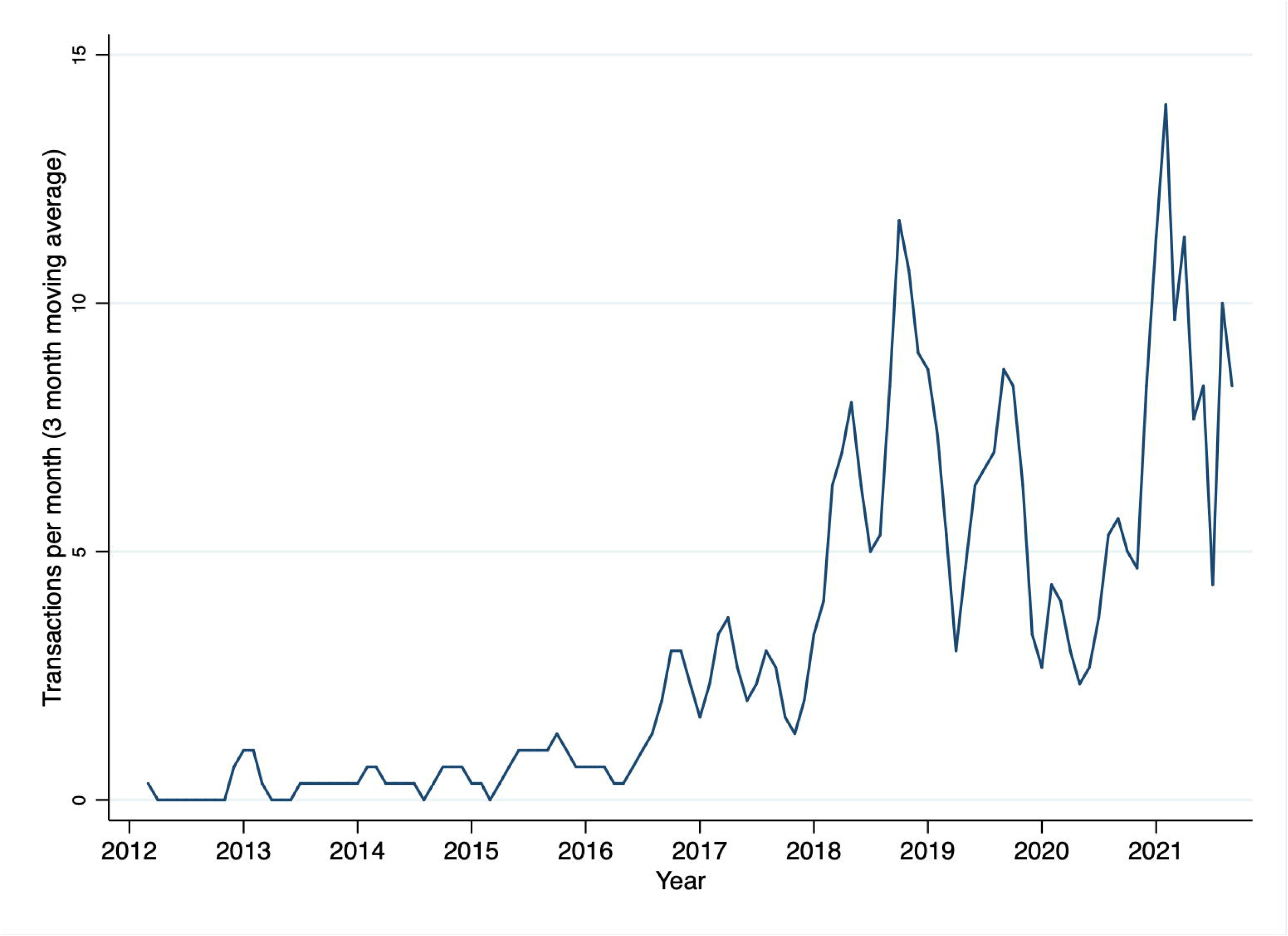
Three month moving average of PE practice acquisitions (transactions) per month, 2012-2021.

**Figure 2:**
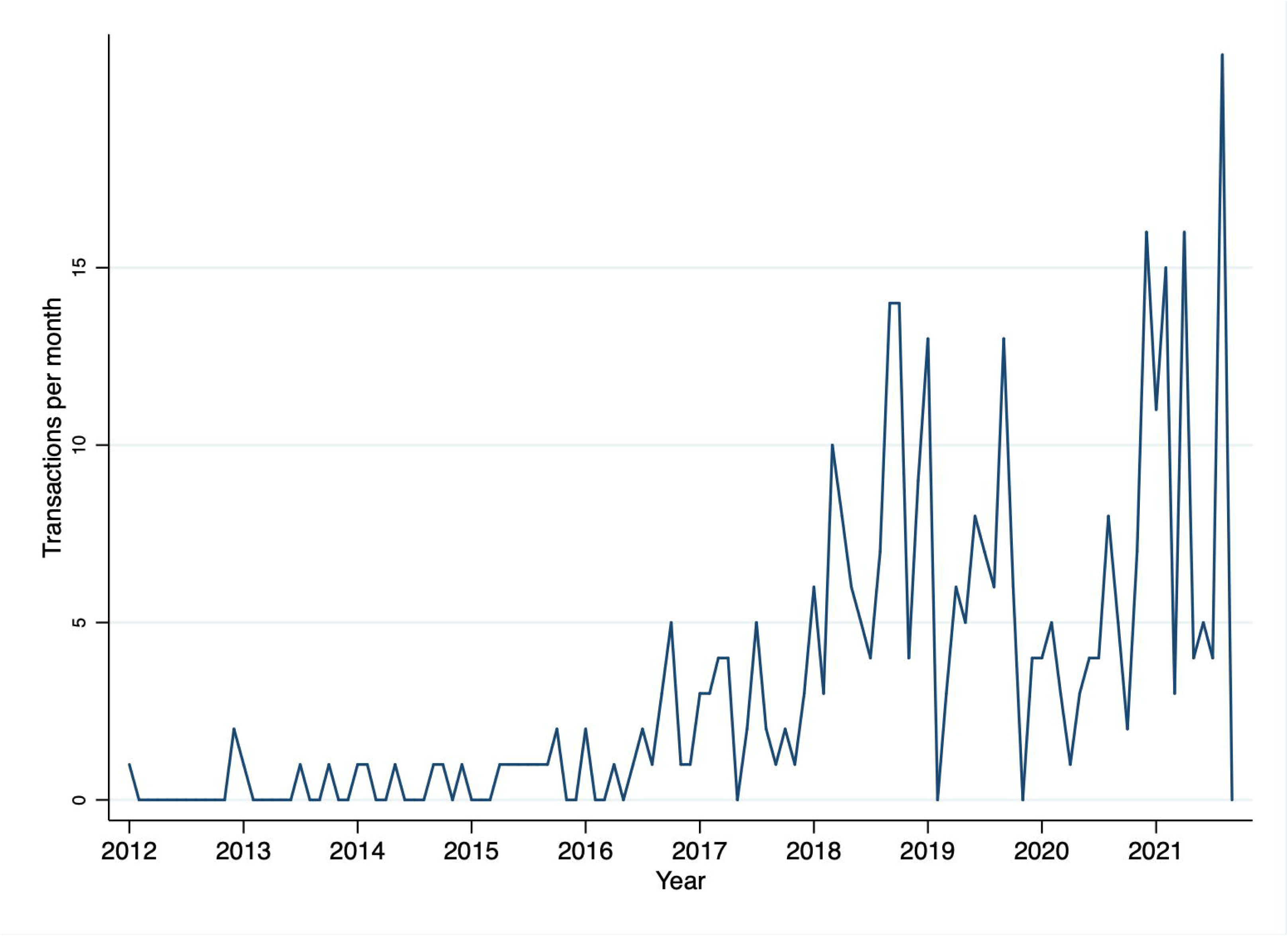
PE practice acquisitions (transactions) per month, 2012-2021.

The average number of acquisitions per month from 1/1/2019-2/29/2020 (pre-COVID) and 1/1/2021-9/1/2021 (COVID post-vaccine) were 5.71/month and 8.78/month (p=0.1971). Average monthly acquisitions from 3/1/2020-12/31/2020 (COVID pre-vaccine) were 5.30/month, similar to that from 1/1/2019 – 2/29/2020 at 5.71/month (pre-COVID) (p=0.8072).

### Financing Status of Platform Companies

One platform company was recapitalized during this time frame, Quigley Eye Specialists, by New Harbor Capital in 2020.

### Geographic Acquisitions

Over the last decade, certain large platform companies with geographic footholds utilized add on acquisitions to expand pre-existing large groups of practices (Figure 3). Texas, Florida, Michigan, New Jersey had larger numbers of acquisitions 55, 48, 29, 28 clinic acquisitions, respectively. Of note, these geographic totals do not include practices for which publicly available information on the clinics acquired could not be found, including AEG vision (50 practices in multiple locations) and Keplr Vision (43 practices in multiple locations).

**Figure 3:**
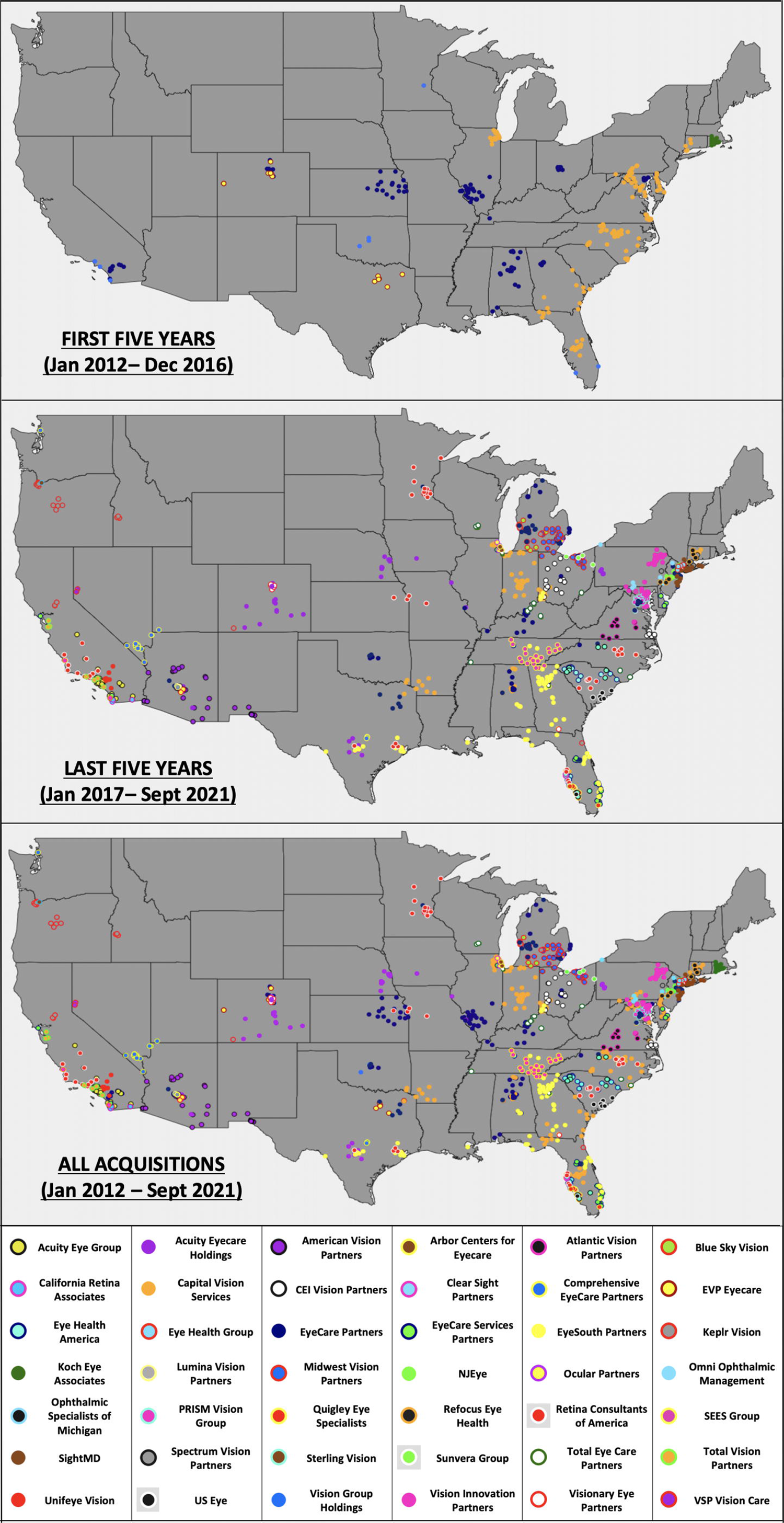
Map showing geographic footprint of ophthalmology and optometry acquisition clinical locations by platform company from January 2012 through December 2016, January 2017 to September 2021, and January 2012 to September 2021.

## Discussion

This study’s findings demonstrate sustained growth of PE backed acquisitions of ophthalmology and optometry practices in recent years throughout the United States, clustered in geographic areas. To characterize this trend, we captured private equity acquisitions of ophthalmology, optometry, and mixed practices starting from 10/21/2019 - 9/1/2021 and compared them to prior data from 1/1/2012 - 10/20/2019. We found that from 2012 to 2021, on average, monthly acquisitions increased by 0.947 acquisitions per year (p<0.001*).

As has been seen in other fields of medicine, private equity acquisitions continue to increase, with certain platform companies serving as hubs in geographic areas (i.e., Midwest Vision Partners, EyeSouth Partners). Particularly, given the nature of ophthalmology as a field of many independent practitioners, we anticipate consistent with previous literature, the PE model specifically targets high value “platform practices,” and then works to acquire add-on practices to scale them up[21]. Consistent with this trend, we found that practices with a median of 3 ophthalmologists were acquired by PE in this time frame.

The ever-increasing presence of private equity in healthcare remains an ethically ambiguous question which will require rigorous discussion and research to fully understand the effects on patients, physicians, and the economy at large. On one hand, consolidation of independent practices in ophthalmology, which is largely consistent of small, independent practices, will allow groups to be represented by economically savvy and robust financial groups. Private equity firms utilize capital from investors with the goal to return a profit by adding value or decreasing overhead. This can be done through tactics aimed at consolidation and efficiency such as outsourcing billing, layoffs, and increasing prices and patient volume[22]. The private equity model introduces the ability to implement best practices along a large network of practices and allows some autonomy in that physicians can still be heavily involved with the logistical operations of the practice[23].

Various elements interplay to make the environment ripe for private equity consolidation. Physicians may be drawn to large buyouts from private equity in the form of large upfront payments from sales of their practice. The large payouts may be taxed at capital gains rates rather than income tax rates, making them particularly appealing[22]. Declining reimbursements for ophthalmologists may also make the large payouts from private equity firms appealing. Additionally, the demand for ophthalmology remains high, and it is predicted to be the surgical subspeciality with the greatest predicted workforce shortage by 2025 [24]. Physicians may opt to merge with private equity due to shared responsibility of financial burdens including the price of expensive equipment[8]. Further, large private equity groups are able to have greater stakes and power during negotiations with insurance companies, which may mitigate the trend of declining reimbursements [8]. Other economic climate factors such as record cash reserves, sophisticated investors, and low interest rates have facilitated a private equity driven marketplace.

In the setting of COVID-19, it was unclear how ophthalmology practices would recover after the necessary curtailing of ophthalmic visits and procedures in light of the Academy of Ophthalmology’s recommendations to professionals[25]. We found that while PE backed acquisitions slowed during the initial phases of the pandemic as depicted graphically, the difference was not statistically significant when comparing the mean number of acquisitions during the COVID-19 pandemic to before the pandemic (i.e., 1/1/2019- 2/29/2020 compared to 3/1/2020 – 12/31 2020 (p=0.8072). Further, when comparing 1/1/2019 - 2/29/2020 to 1/1/2021 to 9/1/2021; or the time before the start of the COVID-19 pandemic compared to after the introduction of the vaccine (p=0.1971) rates were not statistically different, indicating that the introduction of the vaccine may have facilitated sustained growth. The field of ophthalmology has certainly shifted, with a greater emphasis on telemedicine and a keener interest in professional liability insurance and disaster-associated professional services[26]. Given the uncertainty of the state of world affairs and concerns about lockdown orders, PE consolidation would appear attractive to practices with less financial leverage[27].

Despite research indicating that hospitals acquired by private equity were associated with larger increases in net income and charges, the results remain heterogenous as far as patient outcomes[28]. A recent report of private equity investment in nursing homes was associated with increased ambulatory care-sensitive condition emergency department visit, increased hospitalizations, and higher Medicare costs[29]. Further, academic institutions will face new challenges in the setting of private equity expansion, as there will be increased financial strain on centers which traditionally rely on non-aligned general practitioners for subspecialty referrals. This financial strain may have deleterious consequences in terms of the capacity for these academic centers to conduct much of the basic and translational research needed to push the field forward, and to adequately train future residents.

Future ophthalmologists will have to brace for the fact that there will be no mechanism to control their practice, and financial compensation will likely be less with a future revenue stream discounted[30]. Anecdotally, several physicians have written that granular encroachment is not something they have experienced at the practice management level, however, acquisitions are relatively recent and a conflict of interest may be present as many physicians may not be comfortable publicly speaking negatively about their employer [31].

Further concerns regarding PE companies creating local monopolies may also affect both patient and physician choice when seeking care, employment, or referral centers. Previous work outlines a trend of platform companies creating geographic strongholds in certain parts of the United States, which continues to be a trend in this new data[7]. It has been documented that a sizeable number of patients will chose to go to a local hospital independent of decreased mortality risk, higher survival benefit, or revision risk compared to a regional hospital. Other studies have shown that patients do not wish to travel more than 4 hours to obtain the care they need [32–34]. Given that patients will not typically travel large distances for care, it is important to consider that these private equity firms are creating localized monopolies such that patients are limited in the type and location of care they receive. Concerns over private equity creating miniature monopolies has led to calls for anti-trust policymakers from the Federal Trade Commission to take action[35]. We have already seen these concerns in urology as greater than a quarter of urologists in Maryland and New Jersey are employed by PE back companies. In our current study, New Jersey was one of the states with the largest acquisitions with presence in two major metro areas of New York City and Philadelphia.

There are limitations to this study. Two acquisitions were very large and impossible to add to our geographic depiction of acquisitions. This was Keplr Visions acquisition of 43 optometry practices and AEG visions’ acquisition of 50 comprehensive or multi-specialty practices. We anticipate that these large, multi-practice acquisitions would further support our finding that private equity acquisitions are growing with add-on practices in certain geographic strongholds. Further, for some practices, if information was not available on the nature of the practice or staff through web archives at the time of acquisition, thus we opted to utilize information from the date closest to the acquisition.

Acquisitions via private equity decreased during the initial phases of the COVID-19 pandemic but overall remained stable in the COVID pre-vaccine era and continued during the COVID post-vaccine era. Ophthalmology has experienced an increased and sustained continuation of PE acquisitions despite the economic ramifications of COVID-19. The continual growth of private equity during times of hardship to ophthalmology practices raises concerns of patient outcomes, healthcare costs, physician referral choice, and physician employment choice.

## Data Availability

All data produced in the present study are available upon reasonable request to the authors.

## Notes

Conflict of Interest/Financial disclosures Sachi A. Patil: No financial disclosures Daniel G. Vail: No financial disclosures Jacob T. Cox: No financial disclosures. Evan M. Chen: No financial disclosures Prithvi Mruthyunjaya: No financial disclosures James C. Tsai, MD, MBA: Financial Disclosures: Consultant for Eyenovia, ReNetX Bio, Smartlens Ravi Parikh: Financial disclosures: Consultant fees for Anthem, blue cross blue shield

### Competing Interest Statement

James C. Tsai, MD, MBA: Financial Disclosures: Consultant for Eyenovia, ReNetX Bio, Smartlens
Ravi Parikh: Financial disclosures: Consultant fees for Anthem, blue cross blue shield

### Funding Statement

This study did not receive any funding

